# Pulsed X-Ray CT with Temporally Aware Reconstruction for Three-Dimensional Endovascular Guidance: Phantom Feasibility Study

**DOI:** 10.64898/2026.11.18.26368435

**Authors:** Navid Azimi, Arnaud Brian-Choux, Stephen Araujo, Biju Jacob, Eri Haneda, Earl Adamy, Eran Cohen, Simon Sharon, Zhye Yin, Nariman Nezami, Bruno De Man, Amir Pourmorteza

## Abstract

**Background:** Endovascular fluoroscopy provides limited depth information. Projection-level x-ray pulsing and prior-image reconstruction could enable volumetric CT updates from sparse projection data.

**Purpose:** To assess whether pulsed sparse-view CT combined with temporally aware reconstruction could accurately localize robotically manipulated endovascular devices in a phantom.

**Materials and Methods:** We compared true pulsed acquisitions with matched retrospective sparsification (4–180 projections) in a modular phantom. Eight robotically generated device configurations in an anthropomorphic endovascular phantom were scanned with 1248 projections. Data were retrospectively sparsified to 6, 12, 24, 32, and 52 projections and reconstructed using filtered backprojection and a temporally aware prior-image method. Localization errors were measured relative to full-view filtered-backprojection measurements. Analyses were descriptive.

**Results:** Mean CT number differences between true pulsed and matched retrospectively sparsified images were 4.8–6.2 Hounsfield units; corrected standard deviations were 27.5–29.9 Hounsfield units. Sparse-view filtered backprojection obscured device segments at 6 and 12 projections, whereas temporally aware reconstruction depicted both devices at all projection counts. At 6 projections, median centerline, guidewire-tip, and tip-to-vessel-wall errors were 0.82 mm (interquartile range, 0.10 mm), 0.71 mm (0.12 mm), and 0.03 mm (0.28 mm), respectively. All localization errors were below the 0.944-mm reconstructed voxel diagonal.

**Conclusion:** True pulsed and matched retrospectively sparsified acquisitions showed similar CT numbers in a modular phantom. In an anthropomorphic endovascular phantom, temporally aware reconstruction of retrospectively sparsified data localized robotically manipulated devices within one reconstructed voxel diagonal.

## Introduction

Endovascular procedures rely primarily on fluoroscopic guidance, which provides high temporal resolution two-dimensional images but limited depth information and incomplete visualization of the surrounding anatomy (1). Volumetric CT could improve localization of guidewires and catheters relative to vessels and adjacent structures. However, repeated conventional CT acquisitions would increase radiation exposure and interrupt procedural workflow.

Rapid x-ray switching with a gridded-cathode tube permits exposure only at selected projection angles during each gantry rotation, enabling pulsed sparse-view CT (2). Reconstructing a complete image from very few projections with conventional methods produces severe angular undersampling artifacts. Temporally aware reconstruction can address this limitation by using a prior volumetric image to preserve unchanged anatomy while reconstructing interval changes, including guidewire and catheter motion (3). This approach could separate the radiation required to establish anatomy from the radiation required to update device position.

The purpose of this phantom feasibility study was to evaluate whether pulsed sparse-view CT combined with temporally aware reconstruction could provide accurate three-dimensional localization of robotically manipulated endovascular devices. We hypothesized that the method would depict endovascular device motion and provide accurate localization within one reconstructed voxel across a wide range of projection counts.

## Materials and Methods

This study was performed under a sponsored research agreement between GE HealthCare and Emory University and a material transfer agreement with Microbot Medical. Authors who are not employees of or consultants for GE HealthCare or Microbot Medical had complete control of data acquisition, analysis, and publication of results.

### Experimental Design

Two experiments were performed. Experiment 1 evaluated whether retrospective projection sparsification could serve as a surrogate for true pulsed CT acquisition. This validation was necessary because identical guidewire, catheter, vessel, and contrast configurations could not be reproduced across separate conventional and pulsed scans. Experiment 2 then used retrospective sparsification during robotic device manipulation, allowing each sparse reconstruction to be compared directly with a full-view reference acquired at the identical device configuration.

### CT System

All imaging was performed on a clinical Revolution Apex™ CT scanner (GE HealthCare, Waukesha, WI) equipped with a Quantix160™ gridded-cathode x-ray tube. The GE HealthCare team made custom modifications to the system software and firmware to enable microsecond-scale control of x-ray emission (2, 4–6). All scans were performed in axial acquisition mode with 16-cm z-collimation.

### Experiment 1: Validation of Retrospective Sparsification

A 200-mm-diameter modular CT phantom was used to compare true pulsed acquisition with retrospective projection sparsification. The phantom contained 137 removable 2-mL inserts in a three-dimensional printed housing filled with synthetic 20% ballistic gel (Clear Ballistics, Greenville, SC). Inserts contained iodine-, gadolinium-, and tantalum-based contrast materials, calcified plaques, vascular stents, three-dimensional printed airway and vessel inserts, and microrobotic components (7) (Fig 1).

**Figure 1.**
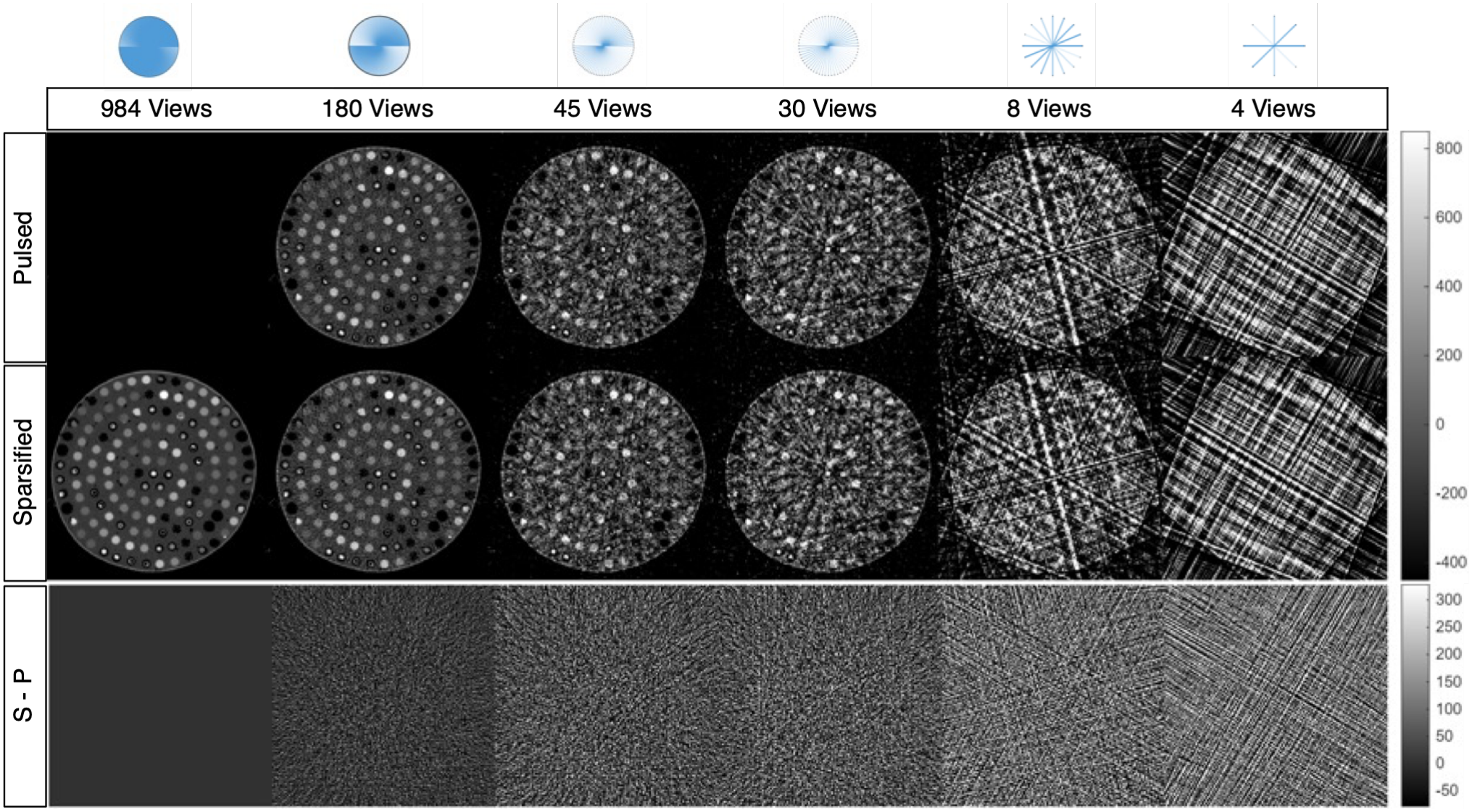
Validation of retrospective sparsification. Angular sampling patterns are shown above. True pulsed acquisitions (top row) used 180, 45, 30, 8, and 4 projections. The conventional 984-projection image and matched retrospectively sparsified images are shown in the middle row. Difference images are shown below.

The phantom was scanned once using a conventional full-view acquisition containing 984 projections. Separate true pulsed scans were then acquired with 180, 45, 30, 8, and 4 projections. All scans used 120 kVp, a 0.28-second gantry rotation, identical acquisition geometry, and a constant exposure per acquired projection (2 mA, 0.56 mAs).

For each pulsed acquisition, projections were selected retrospectively from the conventional 984-projection dataset to match both the number and angular positions of the true pulsed projections within 0.36°.

Agreement was assessed from the voxel-wise difference between each true pulsed reconstruction and its corresponding retrospectively sparsified reconstruction over the entire image volume. The mean difference represented CT number bias. The standard deviation was scaled by the square root of the acquired-to-maximum projection-count ratio to account for differences in total photon count. The result was then divided by 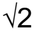 to account for independent noise in the two acquisitions.

### Experiment 2: Robotic Guidewire and Catheter Manipulation

An anthropomorphic endovascular phantom was constructed from a clinical CT angiography dataset (8) with spatial resolution of 0.5 mm isotropic or better. The arterial lumen was segmented in OsiriX (Pixmeo, Geneva, Switzerland), divided into interchangeable anatomic modules, and processed in SolidWorks (Dassault Systèmes, Vélizy-Villacoublay, France) to incorporate vessel-wall thickness and standardized connectors. Vascular components were printed using translucent elastomeric resin (Flexible Tough 2.0, Anycubic, Shenzhen, China). The distal abdominal aorta and iliac artery module was placed within human pelvic bones (Skulls Unlimited International Inc., Oklahoma City, OK) and surrounded by 20% ballistic gel to approximate human x-ray attenuation (Fig 2).

**Figure 2.**
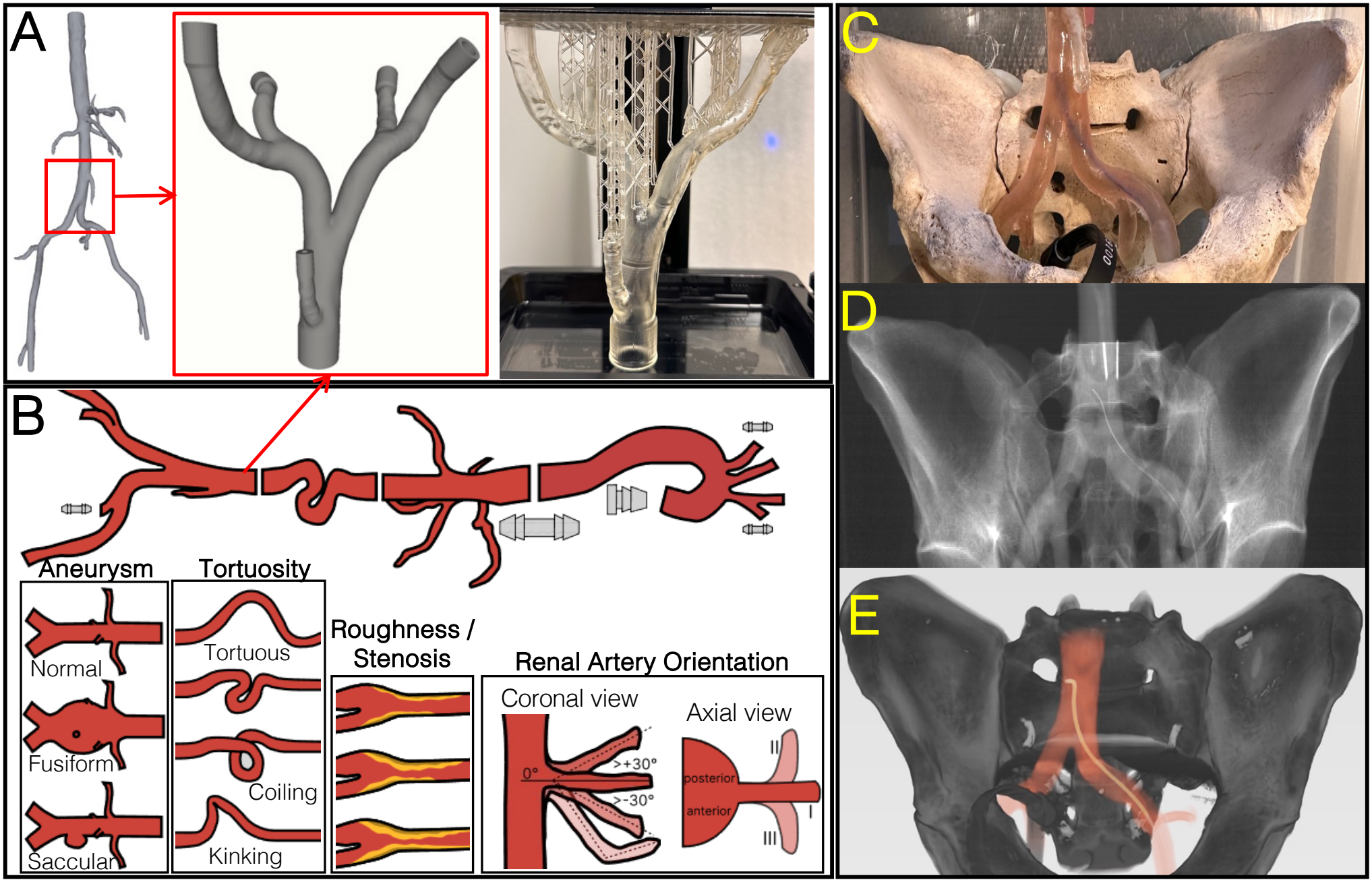
Anthropomorphic endovascular phantom. A. Arterial anatomy segmented from CT angiography, processed in SolidWorks, and fabricated in elastomeric resin. B. Interchangeable vascular modules and connectors of varying anatomic complexity. C. Distal abdominal aorta and iliac artery module positioned within a human pelvis with contrast-filled lumen, microcatheter, and guidewire introduced through the femoral route. D. Fluoroscopic image. E. Three-dimensional volume rendering from a 12-projection Reconstruction of Difference dataset showing bone, vessels, and endovascular devices.

The vascular lumen was filled with iodinated contrast material diluted to 2 mg iodine/mL. A straight tip 2.8 Fr microcatheter and a 0.016-inch flexible-tip guidewire (PROGREAT™ and GLIDEWIRE GT™, Terumo Interventional Systems, Somerset, NJ) were introduced through the femoral access. The devices were guided remotely from the CT control room with the Liberty® endovascular robotic system (Microbot Medical, Hingham, Mass) while the phantom remained within the scanner bore (9).

A full-view reference image acquired before the guidewire entered the imaging field of view was designated as the reference scan. Eight subsequent device configurations were generated robotically through guidewire advancement, retraction, and rotation. One conventional full-view acquisition containing 1248 projections was obtained at each configuration using 70 kVp, 120 mAs, and a gantry rotation time of 1 s. Each full-view dataset was reconstructed with FBP and served as the configuration-specific reference standard.

The corresponding sinograms were retrospectively sparsified to 6, 12, 24, 32, and 52 uniformly distributed projections. Each sparsified dataset was reconstructed with both FBP and Reconstruction of Difference (RoD) (3).

### Image Reconstruction

All datasets were reconstructed offline using the Tomographic Iterative GPU-based Reconstruction toolbox (TIGRE, v2.3) (10) in MATLAB (v2024a, MathWorks, Natick, MA). In Experiment 1, the true pulsed and retrospectively sparsified datasets were reconstructed using filtered backprojection (FBP) only. In Experiment 2, the full-view reference datasets and sparse-view datasets were reconstructed using FBP and RoD. Identical preprocessing and reconstruction settings were used for both experiments, including a Ram-Lak filter and a voxel size of 0.5×0.5×0.625 mm^3^.

RoD is a penalized-likelihood prior-image reconstruction method that estimates changes in sequential imaging protocols from an earlier reference image rather than independently reconstructing the complete image at each time point. The method was implemented as previously described (3). The full-view FBP reconstruction acquired before the guidewire entered the imaging field of view was used as the fixed prior for all reconstructions in Experiment 2. The reconstructed difference volume was added to the grayscale prior image without additional registration.

### Quantitative Device Analysis

Device segmentation was performed using a manually selected fixed CT number threshold that was applied unchanged to all reconstruction conditions and time points. Connected-component filtering was used to remove isolated structures. Within each axial section, the voxel with the highest CT number in the retained device component was selected as the centerline location. The guidewire tip was selected manually. One evaluator with 2 years of CT imaging experience performed the segmentations and measurements. A second evaluator with 18 years of CT imaging experience reviewed and corrected the measurements if necessary.

Three primary localization outcomes were measured. First, geometric agreement of the complete device trajectory was assessed using the standard symmetric Hausdorff distance between the sparse-view RoD and corresponding full-view FBP image centerlines. Second, guidewire-tip localization error was calculated as the three-dimensional Euclidean distance between the tip locations in the sparse-view RoD and full-view FBP reconstructions. Third, tip-to-vessel-wall clearance was calculated as the minimum distance from the guidewire tip to the segmented lumen boundary minus the nominal guidewire radius. Clearance error was calculated relative to the corresponding full-view FBP measurement.

### Statistical Analysis

One conventional acquisition was performed for each device configuration in Experiment 2; all sparsity levels were derived from that acquisition. Separate acquisitions were obtained for each prospectively pulsed sampling condition in Experiment 1. Analyses were therefore descriptive, and no inferential statistical testing was performed. For the eight robotic device configurations, localization measurements were summarized using median, interquartile range, and range, with individual values displayed by projection count. A localization error below the 0.944-mm 3D voxel diagonal was considered within one reconstructed voxel. Tip-to-wall errors were also compared with the 0.5-mm in-plane voxel dimension. Statistical calculations were performed using MATLAB (v2024a, MathWorks, Natick, MA).

## Results

### Validation of Retrospective Sparsification

True pulsed acquisitions were successfully obtained with 180, 45, 30, 8, and 4 projections. Representative pulsed and retrospectively sparsified images showed similar appearance across all projection counts, with minimal residual structure in the difference images (Fig 1) as assessed by a radiologist with more than 10 years of experience. The mean pulsed-minus-retrospective CT number difference was 4.8, 5.2, 4.9, 5.7, and 6.2 Hounsfield units (HU) for 180, 45, 30, 8, and 4 projections, respectively. All mean differences were below 7 HU. The corresponding corrected standard deviations were 28.3, 29.3, 27.5, 28.4, and 29.9 HU. These findings supported the use of retrospective sparsification as a surrogate for true pulsed acquisition in the device-manipulation experiments.

### Sparse-View Reconstruction of Device Motion

Sparse-view FBP produced progressively greater angular undersampling artifacts as the projection count decreased. At 6 and 12 projections, these artifacts obscured portions of the guidewire, catheter, vascular lumen, and surrounding anatomy. In contrast, RoD depicted the guidewire and catheter across all tested projection counts, including 6 and 12 projections (Figs 2E and 3). RoD depicted device advancement, retraction, and rotation across these configurations (Fig 4). RoD images could be added to the prior image to create grayscale images (Fig 3) or superimposed as a colormap on the prior (Fig 4).

**Figure 3.**
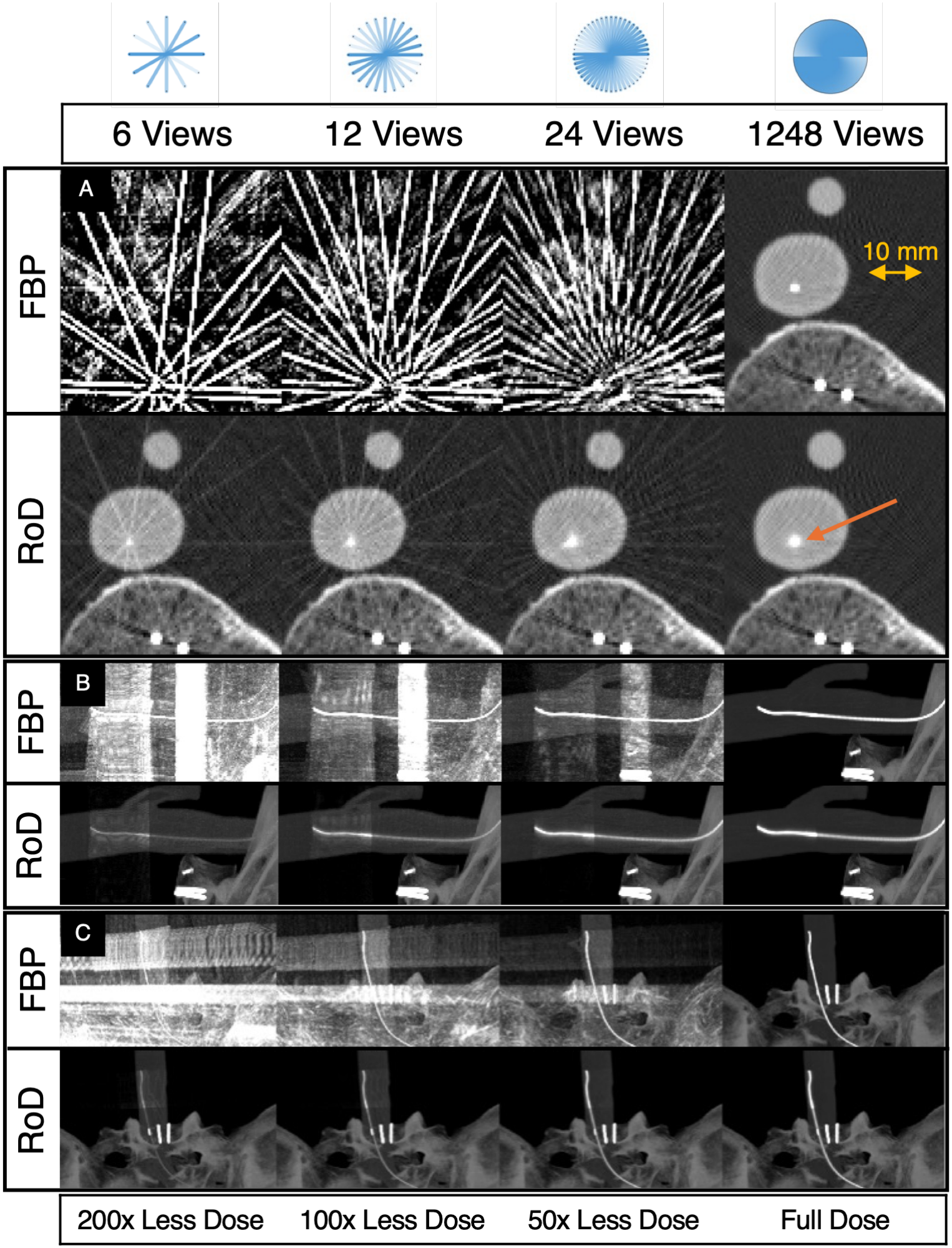
Sparse-view reconstruction of endovascular devices. Columns show reconstructions from 6, 12, 24, and 1248 projections acquired at 70 kVp and 120 mAs. Relative to 1248 projections, the sparse datasets contain 208-, 104-, and 52-fold fewer projections, respectively. These ratios describe potential reductions in exposure for individual update scans at constant exposure per projection; they do not represent measured procedural dose reductions and exclude the full-view prior. A. Axial images; B. sagittal maximum-intensity projections; C. coronal maximum-intensity projections. Sparse-view FBP (upper rows) produced severe undersampling artifacts, whereas Reconstruction of Difference (lower rows) preserved the contrast-filled vessels, microcatheter, and 0.016-inch guidewire, including at 6 and 12 projections. Images were cropped for display from volumes with 160-mm z-axis coverage.

**Figure 4.**
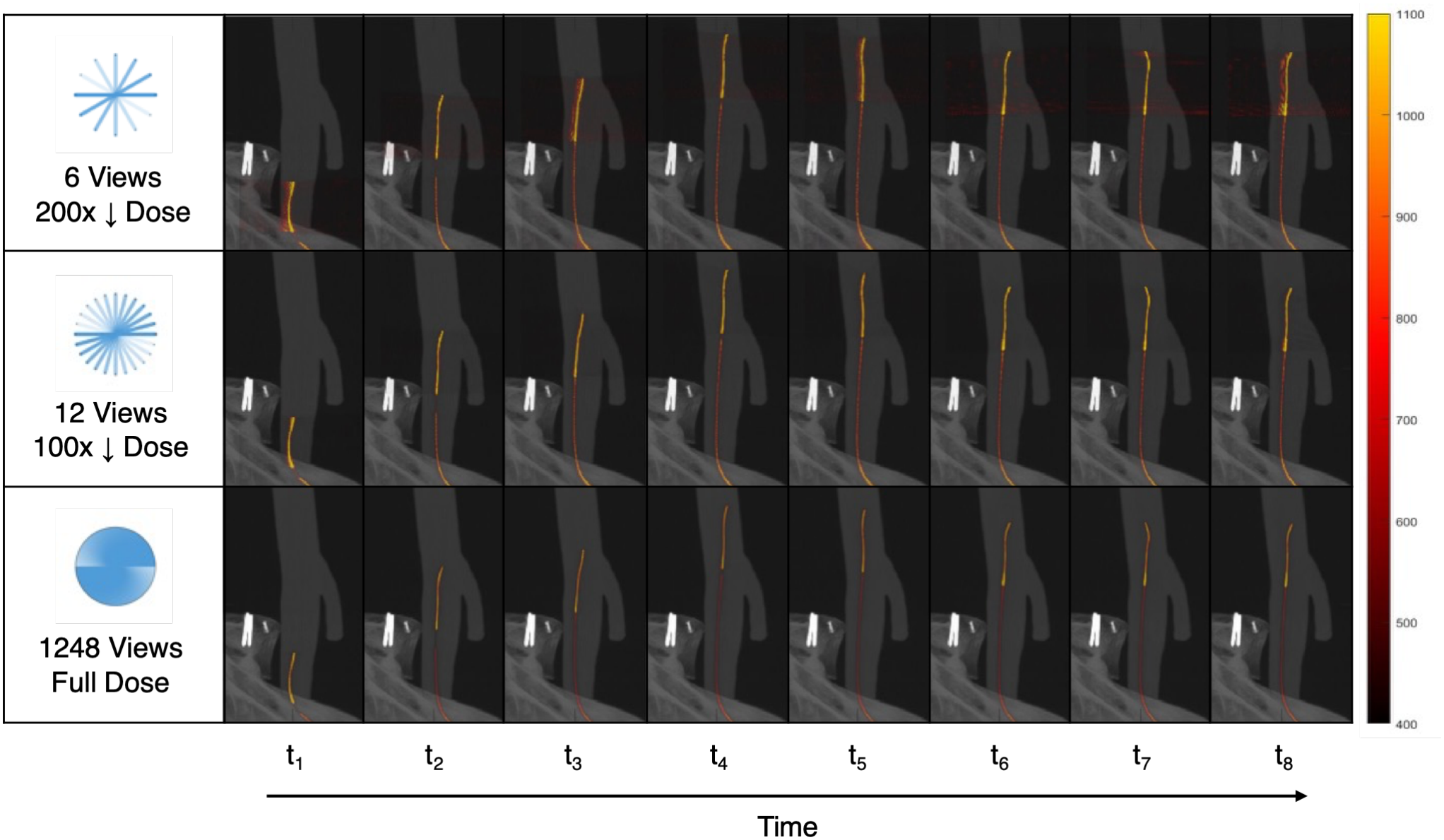
Reconstruction of robotically manipulated endovascular devices. Representative sagittal maximum-intensity projections show guidewire advancement, retraction, and rotation across device configurations. Rows correspond to Reconstruction of Difference images reconstructed from 6, 12, and 1248 projections. Reconstructed device changes are displayed as a color overlay on the fixed grayscale prior image; the color bar indicates CT number in Hounsfield units. Dose labels indicate approximate projection-count ratios for individual update scans and exclude the full-view prior; procedural dose reduction was not measured.

### Device Localization

Across the eight robotic configurations, centerline Hausdorff distances were 0.82 (0.10), 0.59 (0.06), 0.49 (0.03), 0.49 (0.04), 0.50 (0.03), and 0.11 (0.06) mm for 6, 12, 24, 32, 52, and 1248 projections, respectively, reported as median (interquartile range) (Fig 5A). Guidewire-tip localization errors were 0.71 (0.12), 0.53 (0.12), 0.36 (0.07), 0.44 (0.13), 0.45 (0.11), and 0.12 (0.05) mm, respectively (Fig 5B). Tip-to-vessel-wall errors were 0.03 (0.28), −0.05 (0.17), 0.02 (0.09), −0.08 (0.10), 0.05 (0.11), and −0.02 (0.03) mm (Fig 5C). All localization errors were below the 0.944-mm reconstructed voxel diagonal.

**Figure 5.**
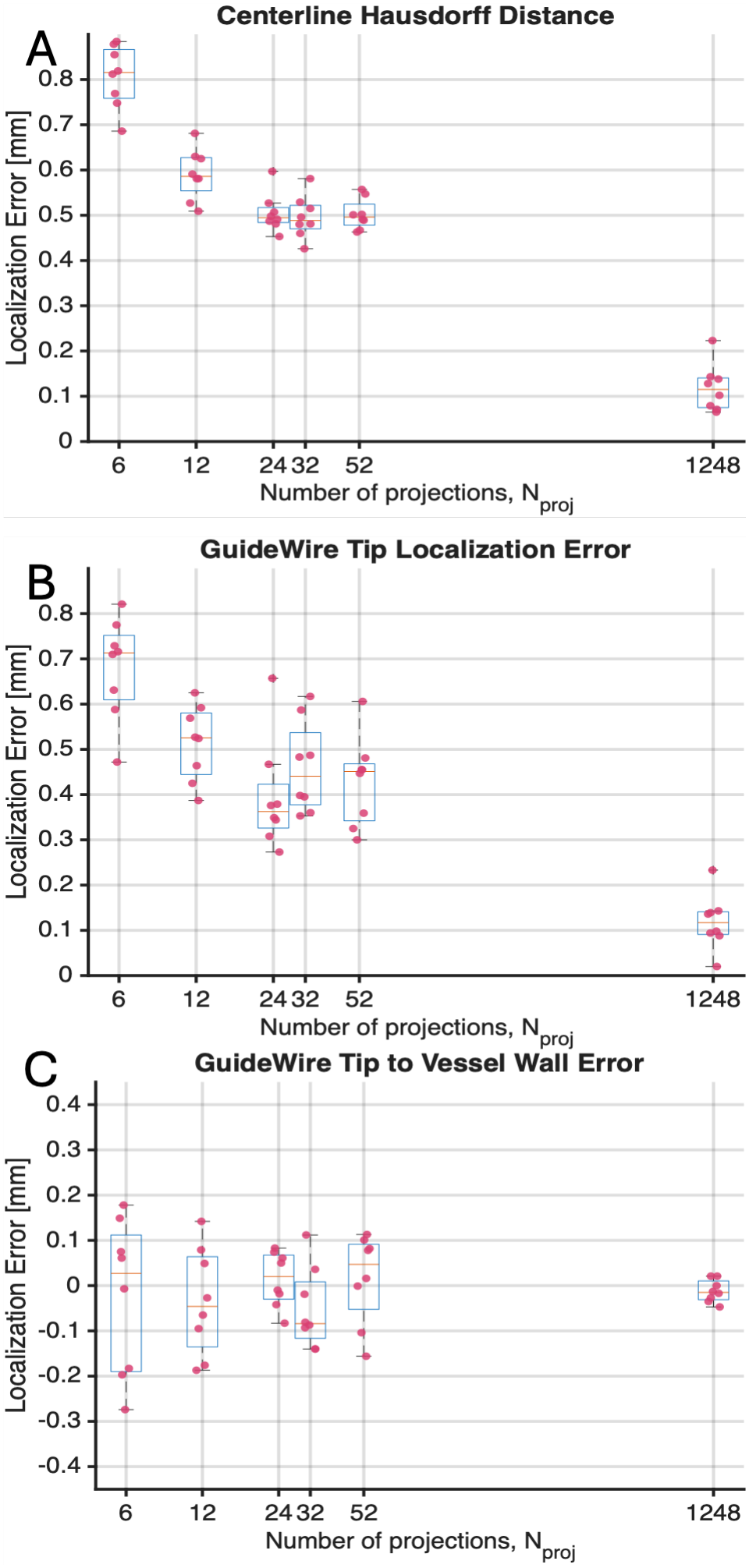
Quantitative device localization across eight robotic configurations. A. Centerline Hausdorff distance; B. guidewire-tip localization error; C. guidewire-tip-to-vessel-wall error for reconstructions using 6, 12, 24, 32, 52, and 1248 projections. Boxes show the interquartile range, center lines show the median, whiskers show the range, and red markers indicate individual measurements. All localization errors were below the 0.944-mm reconstructed voxel diagonal.

## Discussion

We demonstrated true projection-level x-ray pulsing for sparse-view imaging on a clinical CT scanner and validated retrospective sparsification under the tested phantom conditions. Previous sparse-view CT studies have largely relied on retrospective projection removal, while sparse-view cone-beam CT has been limited by slower acquisitions, greater motion sensitivity, flat-panel detector lag, and cone-beam artifacts (11–13). The gridded-cathode tube used here removes a longstanding hardware limitation by allowing x-ray exposure to be controlled at individual projection angles (2). The validation experiment was therefore essential: it established that retrospective sparsification could reasonably represent true pulsed acquisition under the tested conditions and enabled direct comparison of sparse and full-view reconstructions at identical device positions. Related studies using human cardiac CT data demonstrated that sparse pulsed-mode projection sampling retained sufficient information to characterize contrast dynamics and cardiac motion, supporting the feasibility of highly sparse projection acquisition in clinical CT (4, 5).

The principal innovation is the combination of programmable x-ray exposure with temporally aware reconstruction. Pulsing alone reduces the number of exposed projections but leaves conventional FBP vulnerable to severe undersampling artifacts. RoD instead uses a full-view reference to preserve unchanged anatomy and reconstructs only subsequent changes (3). This separates the dose needed to establish anatomy from the dose needed to update device position and provides a general framework for x-ray pulse programming in dynamic CT.

Our findings also demonstrate compatibility with remote manipulation using the LIBERTY® endovascular robotic system. A clinical series has demonstrated the feasibility of this portable, disposable system in peripheral endovascular interventions (9). Remote control may help overcome restricted physical access within the CT gantry and allow the operator to remain outside the scan room during exposure. Acute ischemic stroke is a possible future application. Workflow studies have emphasized reducing imaging-related treatment delays in stroke care (14), but CT-guided thrombectomy would require validation in the relevant anatomy, with clinically appropriate devices and procedural endpoints. This study did not evaluate stroke anatomy or thrombectomy.

The same gridded-cathode tube is also installed on a deep-silicon photon-counting CT system (2, 15). Higher spatial resolution could further improve depiction of small guidewires and catheter tips, while spectral information could improve identification of devices containing high-atomic-number or K-edge materials (7, 16, 17).

This study has four main limitations. First, it used one phantom, scanner, and device combination without physiologic motion or flow. Second, the robotic experiment used retrospective sparsification rather than prospective pulsing. Third, one acquisition was obtained per configuration, and segmentation included manual steps. Fourth, reconstruction was offline; latency, total procedural dose, operator exposure, and comparison with fluoroscopy or cone-beam CT were not evaluated.

Our findings support the feasibility of projection-level x-ray pulsing and temporally aware sparse-view reconstruction for endovascular device localization. The robotic phantom experiment establishes localization accuracy using retrospectively sparsified data; prospective pulsed device guidance, reconstruction latency, and procedural dose remain to be evaluated.

## Abbreviations

CT: computed tomography
FBP: filtered backprojection
FDA: Food and Drug Administration
HU: Hounsfield unit
IQR: interquartile range
RoD: Reconstruction of Difference
TIGRE: Tomographic Iterative GPU-based Reconstruction toolbox

## Declarations

### Funding and sponsor roles

This work was supported by a sponsored research agreement between GE HealthCare and Emory University and a material transfer agreement between Emory University and Microbot Medical Inc. Authors who are not employees of or consultants for GE HealthCare or Microbot Medical had complete control of data acquisition, analysis, and publication of results.

### Ethics approval and consent

Not applicable. This phantom study involved no living participants or identifiable human data.

### Data and code availability

Data and nonproprietary analysis code supporting the findings are available from the corresponding author upon reasonable request, subject to applicable institutional and commercial agreements.

### Prior presentation

Preliminary results were reported in a 2026 conference abstract (6). This manuscript expands that report with validation of true pulsed acquisition, quantitative device-localization analysis across projection counts and robotic configurations, and full methodological details.

## References

1. Eagleton MJ. Updates in Endovascular Procedural Navigation. Canadian Journal of Cardiology 2022;38(5):662–671. doi: 10.1016/j.cjca.2022.02.020

2. Thibault J-B, Utschig M. Revolution Apex with Quantix 160: When Power Meets Coverage. GE Healthcare, 2020.

3. Pourmorteza A, Dang H, Siewerdsen JH, Stayman JW. Reconstruction of difference in sequential CT studies using penalized likelihood estimation. Physics in Medicine and Biology 2016;61(5):1986–2002. doi: 10.1088/0031-9155/61/5/1986

4. Wu P, Haneda E, Pack JD, Jansen IH, Hsiao A, McVeigh E, De Man B. Cardiac Phase Estimation Using Deep Learning Analysis of Pulsed-Mode Projections: Toward Autonomous Cardiac CT Imaging. IEEE transactions on medical imaging 2025;44(6):2424–2435.

5. Haneda E, Heukensfeldt Jansen IM, Wu P, Pack JD, Hsiao A, McVeigh E, De Man B. Bolus tracking from pulsed x-ray projections: A feasibility study using a five-dimensional cardiac CT contrast dynamics model. Medical physics 2025;52(1):131–145.

6. Pourmorteza A, Brian-Choux A, Aguilar O, Yin Z, Haneda E, Jacob B, De Man B, Nezami N. Abstract No. 542 CT Guidance for Intravascular Procedures at 200x Lower Dose via Pulsed X-ray Sparse Acquisition. Journal of Vascular and Interventional Radiology 2026;37(4). doi: 10.1016/j.jvir.2026.108450

7. Brian-Choux AR, Holmes TW, Yin Z, Hoang KB, Salyapongse A, Ansari A, Pourmorteza A. Ultrahigh-resolution K-edge imaging for guidance of microrobots in minimally invasive interventions: A feasibility study. Medical Physics 2025;52(12):e70105. doi: 10.1002/mp.70105

8. Koitka S, Baldini G, Kroll L, van Landeghem N, Haubold J, Kim MS, Kleesiek J, Nensa F, Hosch R. SAROS, A large, heterogeneous, and sparsely annotated segmentation dataset on CT imaging data. Version 1 ed: The Cancer Imaging Archive, 2023.

9. Khoury JK, Lima NJ, Elsahhar A, Li H, Moon JT, Pourmorteza A, Prologo D, Kim J, Bercu Z. Portable endovascular disposable robotic assistant (PEDRA) for image-guided peripheral endovascular interventions: technical feasibility, safety, and future implications. CVIR Endovascular 2026;9(1):141. doi: 10.1186/s42155-026-00789-2

10. Biguri A, Dosanjh M, Hancock S, Soleimani M. TIGRE: a MATLAB-GPU toolbox for CBCT image reconstruction. Biomedical Physics & Engineering Express 2016;2(5):055010. doi: 10.1088/2057-1976/2/5/055010

11. Bian J, Siewerdsen JH, Han X, Sidky EY, Prince JL, Pelizzari CA, Pan X. Evaluation of sparse-view reconstruction from flat-panel-detector cone-beam CT. Physics in Medicine and Biology 2010;55(22):6575–6599. doi: 10.1088/0031-9155/55/22/001

12. Siewerdsen JH, Jaffray DA. A ghost story: spatio-temporal response characteristics of an indirect-detection flat-panel imager. Medical Physics 1999;26(8):1624–1641. doi: 10.1118/1.598657

13. Barral M, Chevallier O, Cornelis FH. Perspectives of Cone-beam Computed Tomography in Interventional Radiology: Techniques for Planning, Guidance, and Monitoring. Techniques in Vascular and Interventional Radiology 2023;26(3):100912. doi: 10.1016/j.tvir.2023.100912

14. Requena M, Olivé-Gadea M, Muchada M, Hernández D, Rubiera M, Boned S, Piñana C, Deck M, García-Tornel Á, Díaz-Silva H, Rodriguez-Villatoro N, Juega J, Rodriguez-Luna D, Pagola J, Molina C, Tomasello A, Ribo M. Direct to Angiography Suite Without Stopping for Computed Tomography Imaging for Patients With Acute Stroke: A Randomized Clinical Trial. JAMA Neurology 2021;78(9):1099–1107. doi: 10.1001/jamaneurol.2021.2385

15. U.S. Food and Drug Administration. Photonova Spectra and Photonova Spectra Select, 510(k) Premarket Notification K253520. Silver Spring, MD: U.S. Food and Drug Administration, 2026.

16. Holmes TW, Yin Z, Stevens GM, Slavic S, Okerlund DR, Maltz JS, Pourmorteza A. Ultra-high-resolution spectral silicon-based photon-counting detector CT for coronary CT angiography: Initial results in a dynamic phantom. Journal of Cardiovascular Computed Tomography 2023;17(5):341–344. doi: 10.1016/j.jcct.2023.08.003

17. Holmes TW, Yin Z, Bujila R, Madhav P, Kim C, Maltz JS, Pourmorteza A. Ultrahigh-Resolution K-Edge Imaging of Coronary Arteries With Prototype Deep-Silicon Photon-Counting CT: Initial Results in Phantoms. Radiology 2024;311(3):e231598. doi: 10.1148/radiol.231598

